# The Effects of External Laser Positioning Systems for MRI Simulation on Image Quality and Quantitative MRI Values

**DOI:** 10.64898/2026.03.06.26347809

**Authors:** Lucas McCullum, Yao Ding, Clifton D. Fuller, Brian A. Taylor

**Affiliations:** UT MD Anderson Cancer Center UTHealth Houston Graduate School of Biomedical Sciences, Houston, USA; Department of Radiation Oncology, The University of Texas MD Anderson Cancer Center, Houston, TX, USA; Department of Radiation Physics, The University of Texas MD Anderson Cancer Center, Houston, TX, USA; Department of Imaging Physics, The University of Texas MD Anderson Cancer Center, Houston, TX, USA

**Keywords:** Magnetic Resonance Imaging, MRI Simulation, External Laser Positioning System, Image Quality, Quantitative Imaging

## Abstract

**Background and Purpose:** Magnetic resonance imaging (MRI) for radiation therapy treatment planning is currently being used in many anatomical sites to better visualize soft tissue landmarks, a technique known as an MRI simulation. A core component of modern MRI simulation configurations are the use of external laser positioning systems (ELPS) to help set up the patient. Though necessary for accurate and reproducible patient setup, the ELPS, if left on during imaging, may interfere negatively with image quality due to leaking electronic noise, of which MRI is sensitive to. It is currently unknown whether this leakage of electronic noise may further affect quantitative values derived from clinically employed relaxometric, diffusion, and fat fraction sequences. Therefore, in this study, we aim to characterize the impact of MRI simulation lasers on general image quality and quantitative imaging accuracy.

**Materials and Methods:** First, a cine acquisition was used to visualize the real-time changes in image signal-to-noise ratio (SNR) from when the ELPS was deactivated to activated. To validate this effect quantitatively, the SNR was measured using the American College of Radiology (ACR) recommended protocol in a homogeneous phantom with the integrated body, 18-channel UltraFlex small, 18-channel UltraFlex large, 32-channel spine, and 16-channel shoulder coils. Next, a geometric distortion algorithm was tested in two vendor-provided phantoms while using the integrated body coil and the ACR Large Phantom protocol was tested. Finally, a series of quantitative MRI scans were performed using a CaliberMRI Model 137 Mini Hybrid phantom to validate quantitative T1, T2, and ADC while a Calimetrix PDFF-R2* phantom was used for quantitative PDFF and R2*. All scans were performed with both the ELPS both deactivated and activated.

**Results:** Visible electronic noise artifacts were seen when using the integrated body coil when the ELPS was activated on the cine acquisition which led to a four-fold decrease in SNR using the ACR protocol. This SNR drop was not seen when using the remaining tested coils. The automatic fiducial detection algorithm was affected negatively by ELPS activation leading to misidentification when identified perfectly with the ELPS deactivated. Degradation in image intensity uniformity, percent signal ghosting, and low contrast object detectability was seen during ACR Large Phantom testing using the 20-channel Head/Neck coil. Concordance across quantitative MRI values was similar when the ELPS was both deactivated and activated while a consistent increase in standard deviation inside the ADC vials was seen when the ELPS was activated.

**Discussion:** The extra noise induced from the activation of the ELPS during imaging should be avoided due to its potential to unnecessarily increase image noise. This is particularly true when conducting mandatory quality assurance testing for image quality and geometric distortion which utilize the integrated body coil which is most susceptible to ELPS-induced noise. Clear clinical guidelines should be implemented to make this issue known to the MRI technologists, physicists, and other relevant staff using an MRI with a supplementary ELPS for patient alignment.

**Graphical Abstract:** 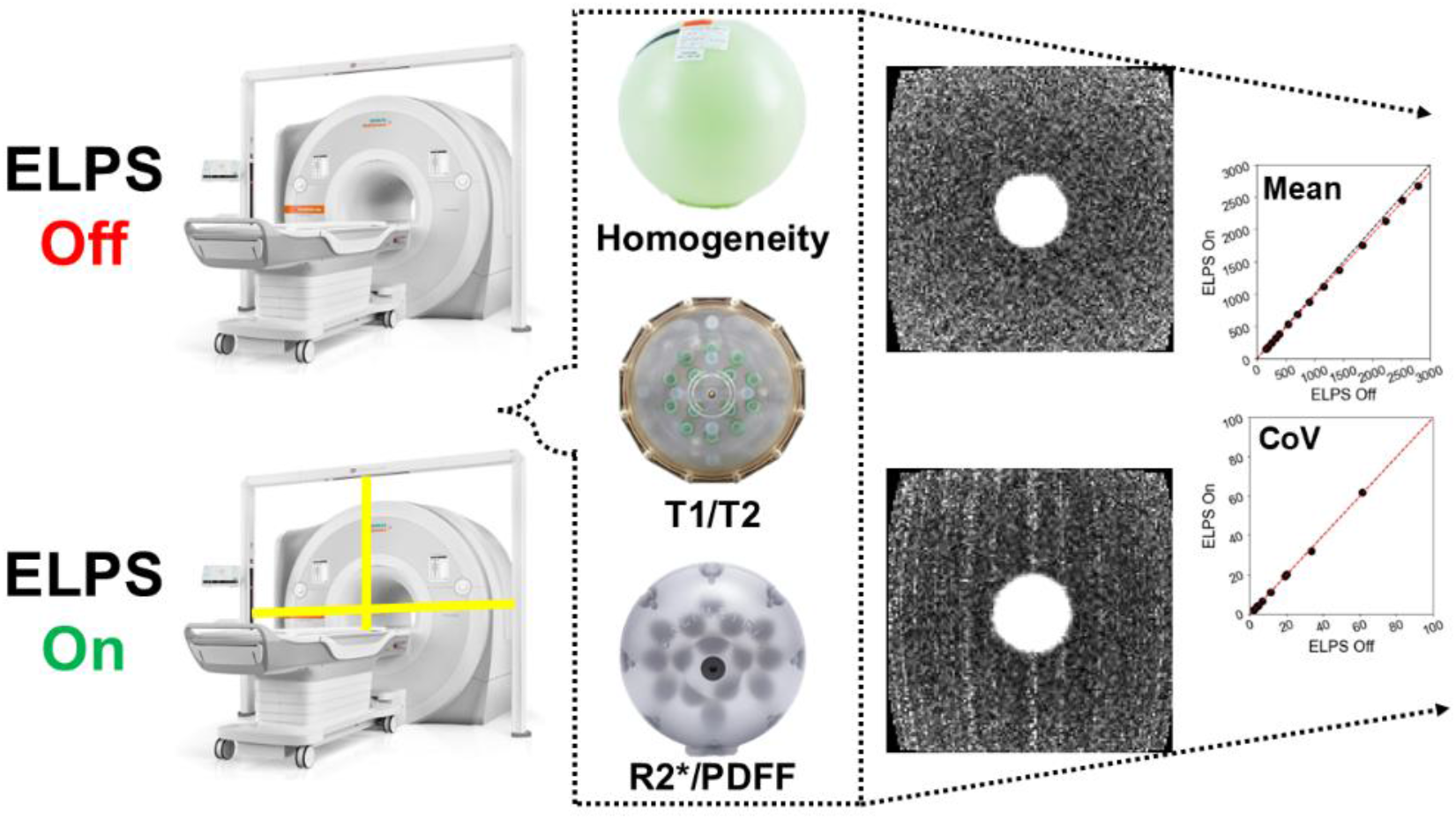

## 1. Introduction

Magnetic resonance imaging (MRI) for radiation therapy treatment planning is currently being used in many anatomical sites to better visualize soft tissue landmarks, a technique known as MRI simulation. A core component of modern MRI simulation configurations are the use of external laser positioning systems (ELPS) to help set up the patient. Though necessary for accurate and reproducible patient setup, these devices, if left on during imaging, may interfere negatively with image quality due to leaking electronic / radiofrequency (RF) noise^1^, of which MRI is sensitive to^2^. These artifacts are commonly referred to as “zipper” artifacts due to their often zipper-like appearance which aligns along the phase encoding direction, or orthogonal to the frequency encoding direction^3^. They should be differentiated from k-space spikes, or “corduroy” / “herringbone” artifact, where stripes are introduced uniformly throughout the whole image with angle and periodicity directly related to the location of the spike in k-space^4^. The frequency of the interference can be approximated by its location in the image along with the receiver bandwidth. Single frequency components will appear as thin lines, while more broadband frequency sources will appear as wider lines or even global image noise if the range of frequencies is large enough. The blips along the phase encoding direction that create the zipper-like appearance are due to the periodicity of the RF interference and can be approximated using the pixel offset of the blips and the repetition time (TR) of the acquisition.

The most common ELPS used in MRI simulation is manufactured by LAP (LAP of America Laser Applications L.L.C.; Boynton Beach, FL, USA). These devices are noted as MR Conditional in the user manual with the requirement that both the laser positioning system and monitor are switched off during MR scanning. This can be done from a master switch which should be included in the clinical workflow of MRI technologists. If this is not done, then reciprocal effects may occur between the LAP system and the MR scanner due to its 50 – 60 Hz operating frequency as noted in the user manual. Contrary to initial thoughts, it is not sufficient to simply turn off the lasers. Instead, the entire system must be deactivated which would deactivate all lights seen on the system including that of the monitor. Therefore, the intended clinical workflow for the system would be to activate it during patient positioning, ensure its deactivation during scanning, and re-activate it for the next patient. However, cumbersome placement of the master switch or improper training of the MRI technologists can lead to scans being conducted while the ELPS is still powered on.

At our institution, we have had instances where quality assurance and periodic maintenance noise tests have failed due to the ELPS being activated, while passing when the ELPS was deactivated again. This unintended discovery led us to notice image quality concerns with deactivation of the ELPS system at our institution during MRI simulation scans due to lack of education and logistical burdens regarding the placement of the deactivation switch. This issue is of primary concern since preventable noise may be incorporated into the images on our MRI simulation scanners. Further, our institution frequently incorporates quantitative imaging including T1 and T2 mapping using multi-dynamic multi-echo (MDME) sequences^5^, apparent diffusion coefficient (ADC) maps^6^ from diffusion-weighted imaging (DWI), and quantitative Dixon producing R2* and proton density fat fraction (PDFF) maps^7,8^ into our scanning workflow and it is currently unclear whether the activation of the ELPS during scanning affects the quantitative values of these scans in terms of bias or repeatability. Another unknown is whether the artifacts are of significant concern only when using the integrated body coil as in the quality assurance and period maintenance tests, or when clinical multi-channel array coils are used as well.

Therefore, the purpose of this paper was to determine: 1) the direct effect of the activated ELPS on image quality through image artifacts and presence of noise, 2) the artifact’s presence across different coil types used, and 3) the subsequent effect of these conditions on quantitative MRI bias and repeatability.

## 2. Methods and Materials

### 2.1. Phantom Assessment

A homogeneous phantom in a container with 1900 mL of “Solution N” (3.75g NiSO_4_ x 6H_2_O and 5g NaCl per 1,000g H_2_O) was used for signal-to-noise ratio (SNR) assessment and artifact identification. The American College of Radiology (ACR) Large Phantom was used to perform the wide range of tests defined in their MRI Accreditation Program. The CaliberMRI Model 137 Mini Hybrid Phantom (CaliberMRI; Boulder, CO, USA) was used as a reference for National Institute of Science and Technology (NIST)-traceable T1, T2, and ADC values. Additionally, the Calimetrix Model 725 PDFF-R2* phantom^9^ (Calimetrix; Madison, WI, USA) was used to validate quantitative R2* and PDFF values. The bore temperature was approximately 20°C across all acquisitions following assessment with an analog thermometer places near the bore. In line with our current clinical workflow for head and neck cancer patients, two 18-channel UltraFlex small and spine coil elements 1 – 3 were used.

Additionally, ACR localizer images were acquired to test the effects of different coils (integrated body coil, 20-channel BioMatrix Head/Neck Tiltable CoilShim – TCS, 18-channel UltraFlex small, 18-channel UltraFlex large, 32-channel spine coil elements 1 – 4, and 16-channel shoulder) on the presence of the RF interference artifact. To determine the extent of the ELPS-induced RF interference artifact, individual spine coil elements were imaged using an ACR localizer while only one element was active. The homogeneous phantom was placed between spine coil elements 1 – 3 for each image acquisition. Since our institution measures geometric distortion using the integrated body coil, we decided to test that workflow with the ELPS both deactivated and activated in two separate phantoms. The first was a vendor-provided 3D geometric distortion phantom from the 1.5T MRI (Philips Healthcare; Best, Netherlands) Linear accelerator (Unity; Elekta AB; Stockholm, Sweden). This phantom has a size of 500 x 375 x 330 mm^3^ with seven 55 mm separated planes along the superior-inferior direction with fiducials in each plane separated by 25 mm. The second phantom was the QUASAR™ GRID^3D^ Image Distortion Phantom (Modus Medical Devices; London, Ontario, Canada) which is a smaller phantom of 152 x 182 x 190 mm^3^ consisting of 1765 control points and accompanying vendor-provided automated geometric distortion analysis.

### 2.2. MRI Acquisition Parameters

All scans were performed on a 3T Siemens Vida (XA60; Siemens Healthcare; Erlangen, Germany) MRI-simulation scanner equipped with an ELPS (DORADOnova MR3T; LAP of America Laser Applications L.L.C.; Boynton Beach, FL, USA). For the homogeneous phantom, a standard phase map was acquired with gradient echo sequence with a field-of-view = 320 x 320 x 250 mm^3^, matrix size = 128 x 128 x 25, reconstructed voxel size = 2.5 x 2.5 x 10 mm^3^, flip angle = 45 degrees, TR = 500 ms, TE = 5 ms, receive bandwidth = 250 Hz/Px, number of signal averages (NSA) = 2, and scan time = 2 minutes 7 seconds. Cine images to assess real-time changes in image quality during ELPS activation were acquired using fast imaging with steady-state precession (TrueFISP^10,11^) sequence with a field-of-view = 256 x 256 x 15 mm^3^, matrix size = 128 x 128 x 1, reconstructed voxel size = 1 x 1 x 15 mm^3^, flip angle = 29 degrees, TR = 160 ms, TE = 1 ms, GeneRalized Autocalibrating Partially Parallel Acquisitions (GRAPPA^12^) factor = 2, receive bandwidth = 1502 Hz/Px, number of dynamics = 513, and scan time = 1 minute 6 seconds. To image the MR-Linac geometric distortion phantom, we developed a T1-weighted fast low-angle shot (FLASH^13^) sequence with FOV = 539 x 539 x 360 mm^3^, matrix = 1088 x 1088 x 180, distance factor = 20%, reconstructed voxel size = 0.5 x 0.5 x 2 mm^3^, TR = 5.03 ms, TE = 2.46 ms, flip angle = 20 degrees, receive bandwidth = 510 Hz/Px, and scan time = 7 minutes 27 seconds. A second T1-weighted FLASH sequence was applied to image the QUASAR geometric distortion phantom with FOV = 256 x 256 x 160 mm^3^, matrix = 256 x 256 x 160, reconstructed voxel size = 1 x 1 x 1 mm^3^, TR = 6.2 ms, TE = 2.7 ms, flip angle = 20 degrees, receive bandwidth = 399 Hz/Px, number of averages = 2, and scan time = 4 minutes 25 seconds. The standard ACR T1-weigthed and T2-weighted sequences as described in the MRI Accreditation Program manual were used to perform the tests on the ACR Large Phantom using the 20-channel Head/Neck coil.

For this study, three clinically deployed quantitative imaging techniques were tested to determine the impact of scanning during ELPS activation on their accuracy and repeatability. First, for quantitative T1 and T2 relaxometry mapping, a multi-dynamic multi-echo (MDME) sequence^14^ was acquired. Second, for quantitative ADC values, a motion correction with radial blades (BLADE) DWI sequence^15^ was used. Finally, a quantitative Dixon (q-Dixon) sequence was used to determine quantitative R2* and PDFF. These sequences were acquired using two 18-channel UltraFlex small coils anterior to the phantom and spine coil elements 1 – 4 posterior to the phantom with detailed acquisition parameters shown in **Table 1**. The repeatability of the quantitative values was determined by conducting five repetitions of each quantitative MRI sequence using their respective phantom (MDME and BLADE DWI for the CaliberMRI phantom and q-Dixon for the Calimetrix phantom) while the ELPS was both activated and deactivated.

**Table 1.** The acquisition parameters for the quantitative MRI sequences used in this study. Note that some parameters may not be valid for the specified sequence which is indicated by a dashed line. Abbreviations: ETL = echo train length, GRAPPA = GeneRalized Autocalibrating Partially Parallel Acquisitions, TE = echo time, TR = repetition time.

|  | MDME | BLADE DWI | q-Dixon |
| --- | --- | --- | --- |
| Field-of-View (mm<sup>3</sup>) | 256 x 256 x 200 | 256 x 256 x 208 | 256 x 256 x 248 |
| Matrix Size | 256 x 256 x 50 | 128 x 128 x 52 | 128 x 128 x 124 |
| Reconstructed Voxel Size (mm<sup>3</sup>) | 1 x 1 x 3 | 2 x 2 x 4 | 2 x 2 x 2 |
| Flip Angle (°) | 150 | 110 | 4 |
| TR (ms) | 7310 | 5900 | 7.95 |
| TE (ms) | 19 / 94 | 38 | 1.03 / 2.15 / 3.27 / 4.39 / 5.51 / 6.63 |
| GRAPPA Factor | 3 | 3 | - |
| Receive Bandwidth (Hz/Px) | 210 | 651 | 1116 |
| Additional Notes | ETL = 12, N<sub>dynamics</sub> = 4, Slice Gap = 0.3 mm | b-values = 0 / 800 s/mm<sup>2</sup>, ETL = 18 | - |
| Scan Time (m:ss) | 5:31 | 5:50 | 3:03 |

### 2.3. Post-Processing and Statistical Analysis

All relevant analysis concerning statistical methods were formulated using the guidelines for reporting Statistical Analyses and Methods in the Published Literature (SAMPL^16^) and all computations unless otherwise specified were performed using Python 3.12.10. The image SNR was determined using Method 4 from the National Electrical Manufacturers Association (NEMA) Standard Publication MS 1-2008. Specifically, this was done by determining the ratio of the mean signal region-of-interest (ROI) in the homogeneous region and the corner noise ROIs with a Gaussian to Rayleigh distribution correction^17^. Quantitative SNR determination was made for the different coil combination images using ImageJ^18^. All ACR Large Phantom analysis was done following the ACR MRI Accreditation Program guidelines from October 19, 2022. The quantitative T1 and T2 values from the MDME were reconstructed using SyMRI StandAlone 11.3.11 (SyntheticMR AB; Linkoping, Sweden) while the ADC, PDFF, and R2* images were taken directly from the scanner reconstruction. For all the quantitative MRI sequences, a region-of-interest (ROI) was created for each vial of the respective phantom with a slight margin to avoid the inclusion of ringing artifacts. All ROIs were created in 3D Slicer^19^ (https://www.slicer.org) and exported in NIfTI format^20^. The fiducials inside the geometric distortion phantom were automatically detected using a custom algorithm using the generalized Hough transform^21–23^. After extracting the quantitative imaging values from each vial of the phantom, Lin’s Concordance Correlation Coefficient (LCCC^24^) was used to assess direct agreement between the mean measured vial values and the phantom reference values. Next, Bland-Altman plots^25^ were used to assess quantitative values that exceeded an expected 95% limit of agreement threshold. A linear fit was computed for reference acquisitions against nominal phantom reference values. Additionally, the standard deviation and subsequent coefficient-of-variation (CoV) inside each vial was computed.

## 3. Results

### 3.1. Non-Quantitative Imaging Analysis

The progression of the RF interference artifact in the cine acquisition is shown in **Figure 1**. Initially, no artifact was seen in **Figure 1(a)**. However, once the ELPS was activated, a broadband RF artifact was seen to the left of the phantom **Figure 1(b)** and gradually shifts towards the left throughout each dynamic of the cine in **Figure 1(c)** and **Figure 1(d)**, effectively reducing the SNR by half. After full activation and when the RF band begins to wrap around, only skinnier lines with blips are seen throughout the image in **Figure 1(e)** indicative of a narrower RF band. Finally, after wrapping around, a brief and subtle artifact similar to **Figure 1(d)** was seen on the right side of the image in **Figure 1(f)** before returning to **Figure 1(a)** once the ELPS has been deactivated.

**Figure 1.**
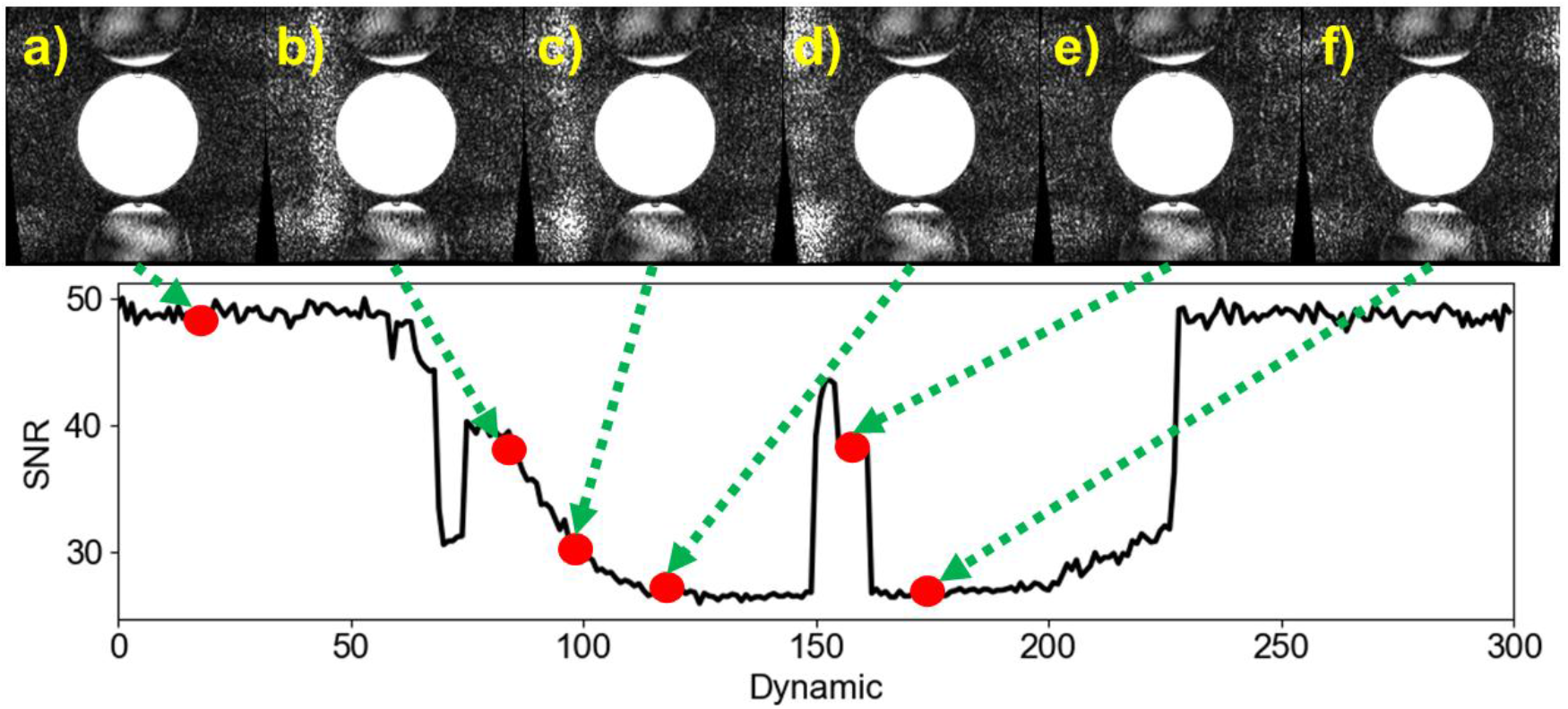
The presentation of the RF interference artifact and its respective SNR change induced by the activation of the ELPS throughout the cine acquisition.

To determine the effect of coil choice, the SNR in the homogenous phantom when the ELPS was deactivated and compared to when it was activated across different coil types is shown in **Table 2**. The integrated body coil was affected the most with an SNR drop from 73 to 19 while the rest of the coil combinations tested showed little difference.

**Table 2.**
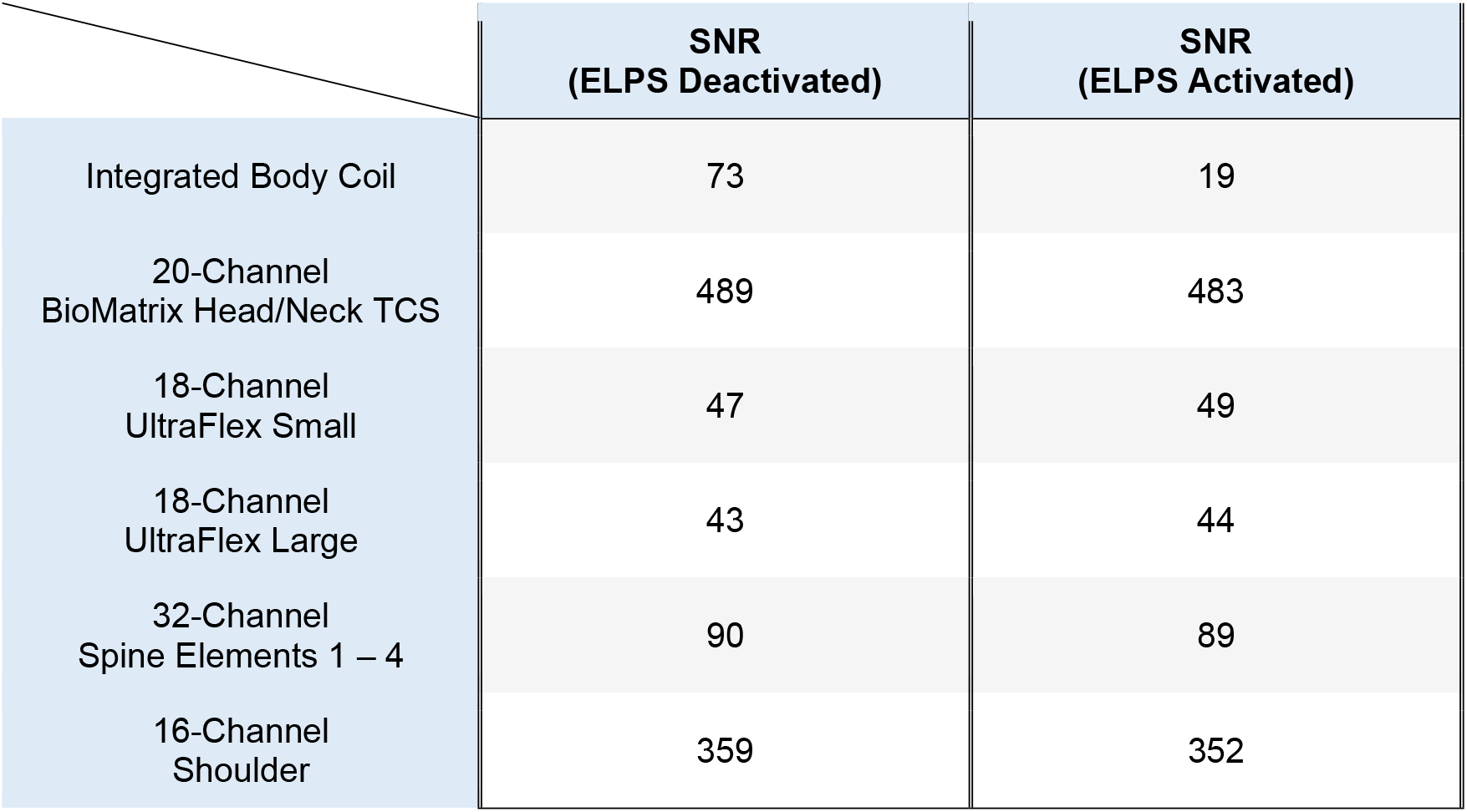
A demonstration of the change in SNR when the ELPS was deactivated and activated in a homogeneous phantom across different coil types.

|  | SNR<br>(ELPS Deactivated) | SNR<br>(ELPS Activated) |
| --- | --- | --- |
| Integrated Body Coil | 73 | 19 |
| 20-Channel<br>BioMatrix Head/Neck TCS | 489 | 483 |
| 18-Channel<br>UltraFlex Small | 47 | 49 |
| 18-Channel<br>UltraFlex Large | 43 | 44 |
| 32-Channel<br>Spine Elements 1 – 4 | 90 | 89 |
| 16-Channel<br>Shoulder | 359 | 352 |

The results of the ACR MRI Accreditation Program tests with both the ELPS deactivate and activated are shown in **Table 3**. A 5% decrease in the image intensity uniformity in the ACR T2-weighted sequence when the ELPS was activated was seen, though a slight increase (91.19 vs. 90.67%) was seen in the ACR T1-weighted sequence. The percent signal ghosting also increased by a factor of 4.5 when the ELPS was activated leading to a failed low contrast object detectability test for the ACR T2-weighted sequence.

**Table 3.** The ACR MRI Accreditation Program test results when the ELPS was deactivated and activated using the ACR Large Phantom. Abbreviations: Ax = axial, Diag = diagonal, LR = lower right, Sag = sagittal, UL = upper left.

|  | ELPS Deactivated | ELPS Activated |
| --- | --- | --- |
| Geometric<br>Accuracy<br>(148 ± 3 mm,<br>190 ± 3 mm) | **Sag Height:** 146.5 mm<br>**Ax Slice 1 Height:** 189.5 mm<br>**Ax Slice 1 Width:** 189.5 mm<br>**Ax Slice 5 Height:** 189.5 mm | **Sag Height:** 145.2 mm<br>**Ax Slice 1 Height:** 189.8 mm<br>**Ax Slice 1 Width:** 189.5 mm<br>**Ax Slice 5 Height:** 189.8 mm |

|  | **Ax Slice 5 Width: 188.8 mm**<br>**Ax Slice 5 Diag Down: 188.9 mm**<br>**Ax Slice 5 Diag Up: 189.0 mm** | **Ax Slice 5 Width: 189.2 mm**<br>**Ax Slice 5 Diag Down: 188.5 mm**<br>**Ax Slice 5 Diag Up: 188.3 mm** |
| --- | --- | --- |
| High Contrast Spatial Resolution ($\leq 1$ mm) | **T1 UL: 1 mm**<br>**T1 LR: 1 mm**<br>**T2 UL: 1 mm**<br>**T2 LR: 1 mm** | **T1 UL: 1 mm**<br>**T1 LR: 1 mm**<br>**T2 UL: 1 mm**<br>**T2 LR: 1 mm** |
| Slice Thickness Accuracy ($5 \pm 1$ mm) | **T1: 5.01**<br>**T2: 5.07** | **T1: 4.59**<br>**T2: 4.88** |
| Slice Position Accuracy ($\leq 7$ mm) | **T1 Slice 1: -1.95**<br>**T1 Slice 11: -1.06**<br>**T2 Slice 1: -2.02**<br>**T2 Slice 11: -0.80** | **T1 Slice 1: 4.15**<br>**T1 Slice 11: 3.91**<br>**T2 Slice 1: 3.23**<br>**T2 Slice 11: 3.85** |
| Image Intensity Uniformity ($\geq 85\%$) | **T1: 90.67%**<br>**T2: 95.56%** | **T1: 91.19%**<br>**T2: 90.88%** |
| Percent Signal Ghosting ($\leq 3\%$) | 0.0038% | 0.017% |
| Low Contrast Object Detectability ($\geq 37$) | **T1: 39**<br>**T2: 37** | **T1: 40**<br>**T2: 35** |

The ACR localizer imaged at each individual spine coil element can be seen in **Figure 2**. As shown, every coil element experiences some form of RF interference artifact from the ELPS system regardless of their location from the ELPS source.

**Figure 2.**
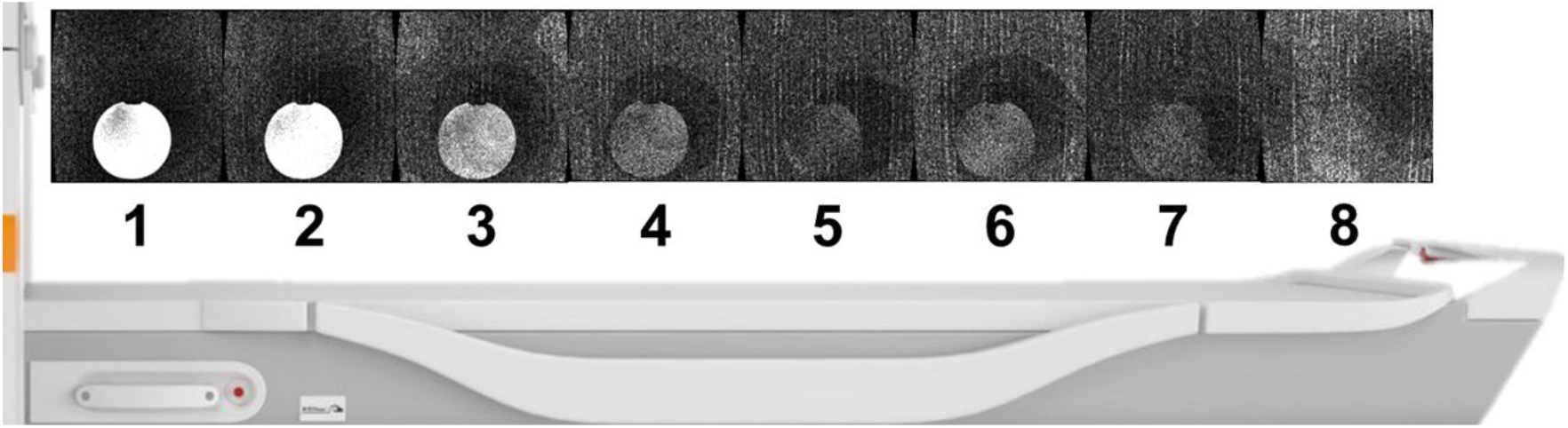
Demonstration of the image quality from each spine coil element of a long homogeneous phantom placed on spine coil elements 1 – 3 while the ELPS was activated. Note, each image uses the same window / level settings for a fair comparison.

A demonstration of the performance of the fiducial location detection in the MR-Linac vendor-provided geometric distortion phantom is shown in **Figure 3**. Note how when the ELPS was activated, more fiducial markers are missed as a false negative due to the increased noise. This is particularly true near the ELPS-induced noise band near the superior portion of the phantom as well as on the inferior edge where clear streaking can be seen from the RF interference.

**Figure 3.**
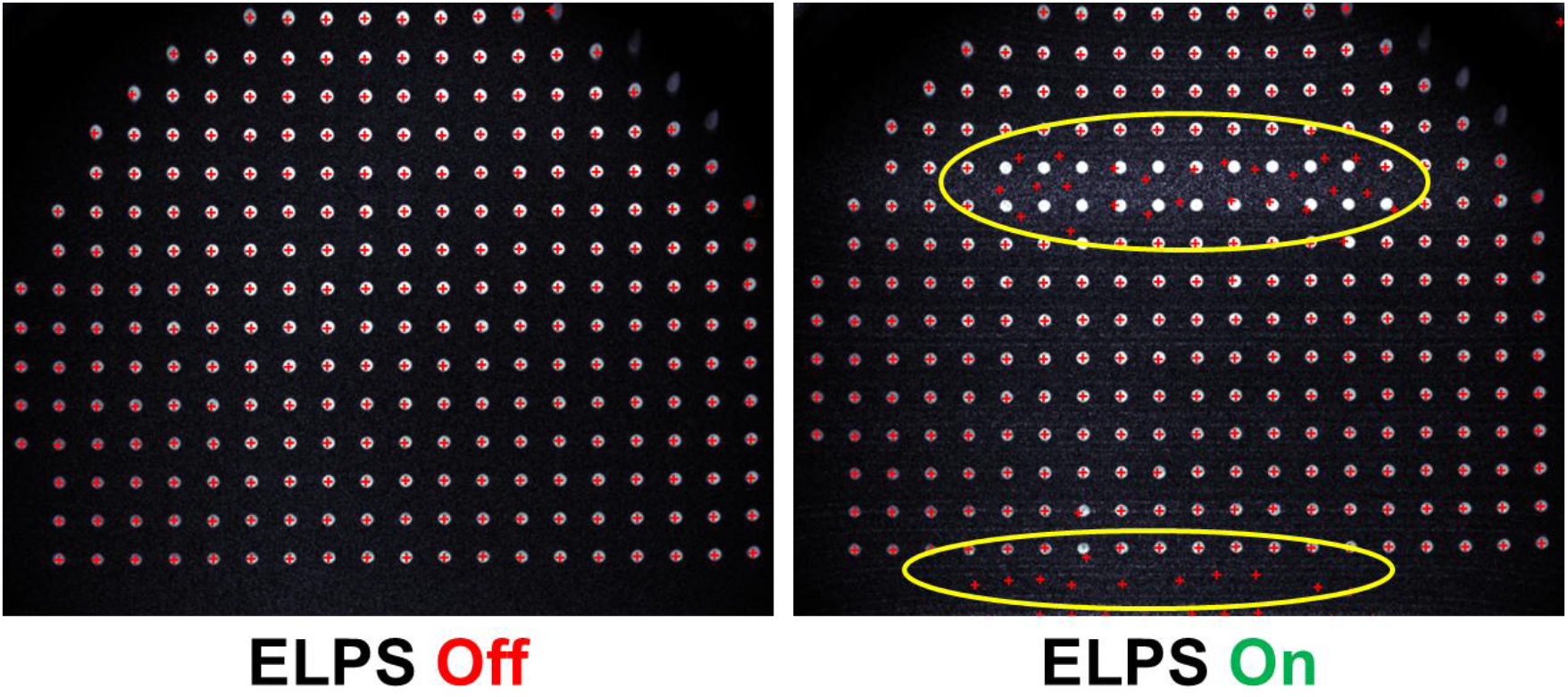
Failure of fiducial recognition from the geometric distortion phantom when the ELPS is activated due to increased background noise while using the body coil obfuscating the fiducial edges. The regions with the largest failures are shown using the yellow circles, particularly the inferior edge and region with the largest RF interference noise band artifact.

A demonstration of the image quality in the QUASAR geometric distortion phantom is shown in **Figure 4**. Note how when the ELPS was activated, an RF interference noise band was seen on the left side of the image as well as on both the inferior and superior edges.

**Figure 4.**
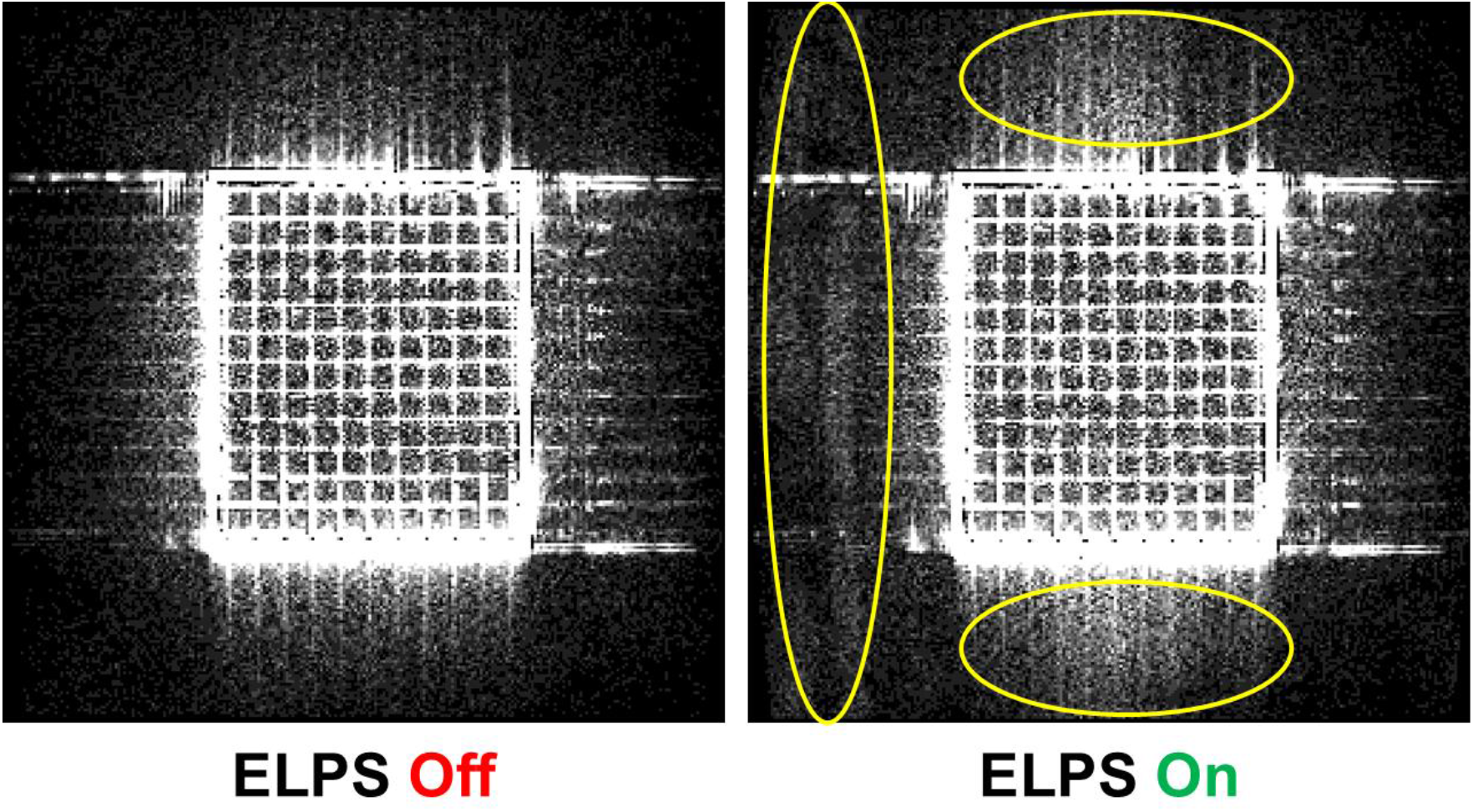
An RF interference noise band artifact is seen with a low window/level setting on the left side of the image while increased general noise is noted in both the interior and superior regions.

The quantitative geometric distortion analysis results from the vendor-provided software is shown in **Table 4**. No substantial differences were noted between when the ELPS was deactivated compared to when it was activated.

**Table 4.** The results of the QUASAR analysis when the ELPS was both deactivated and activated.

|  | ELPS Deactivated | ELPS Activated |
| --- | --- | --- |
| Absolute Distortion Mean (mm) | 0.474 | 0.469 |
| Absolute Distortion Standard Deviation (mm) | 0.143 | 0.142 |
| Absolute Distortion Maximum (mm) | 0.957 | 0.954 |
| Absolute Distortion Percent Above 0.8 mm (%) | 3.147 | 3.247 |

### 3.2. Quantitative Imaging Analysis

Plots of the agreement between the measured mean values and reference values for the quantitative T1, T2, ADC, PDFF, and R2* for when the ELPS was both deactivated and activated are shown in **Figure 5**. Overall, there was minimal difference between quantitative measurements in terms of the linear line-of-best-fit and the resulting LCCC when the ELPS is activated compared to when it was deactivated, however this could be due to the consolidation of the values by using the mean. Therefore, the next plots in this section will give a more detailed analysis of the results.

**Figure 5.**
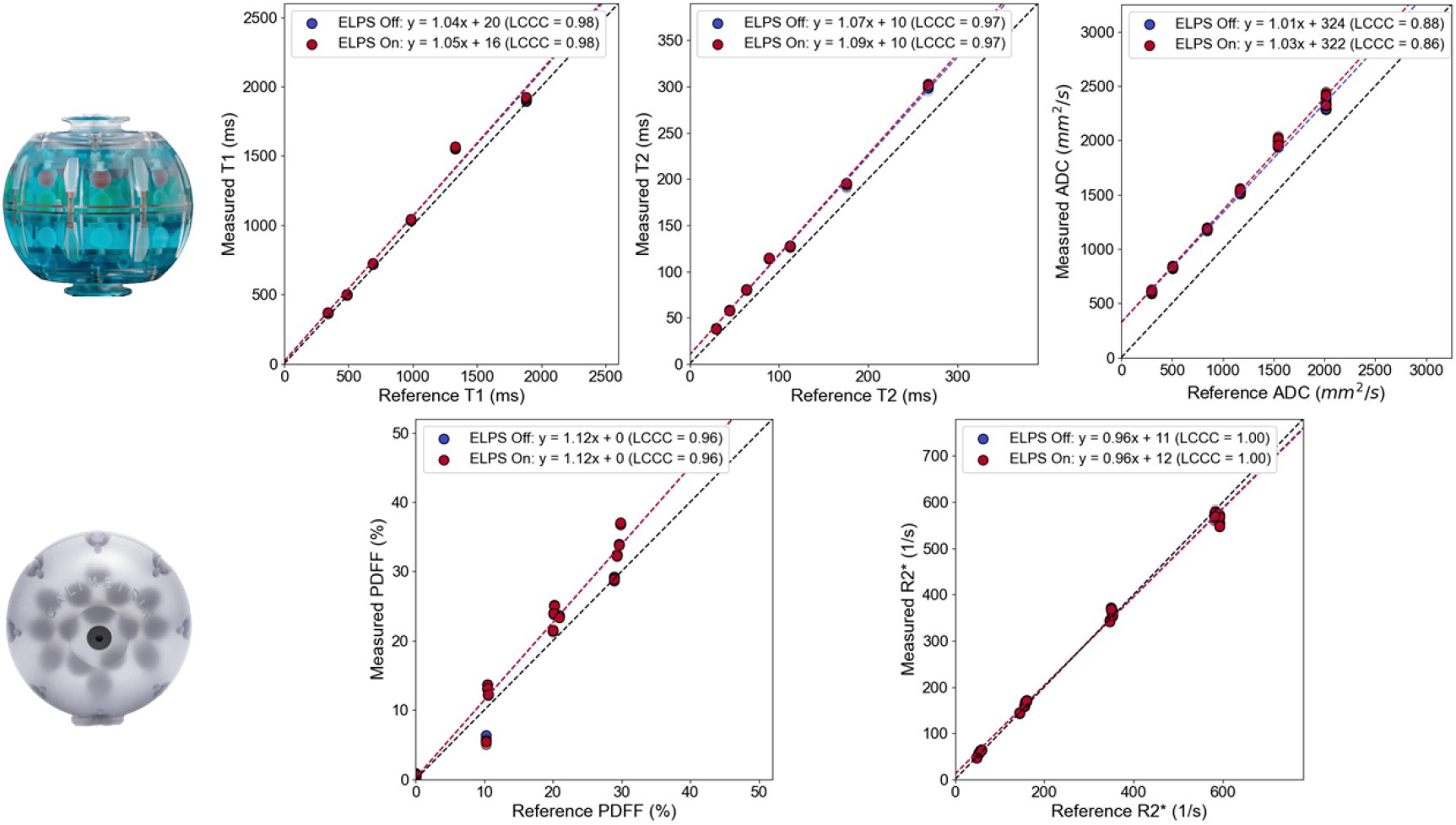
Plots of the agreement between the quantitative T1 (top left), T2 (top middle), ADC (top right), PDFF (bottom left), and R2* (bottom right) with corresponding linear lines-of-best-fit and LCCC values for when the ELPS is both deactivated (blue) and activated (red).

The respective Bland-Altman plot for **Figure 5** is shown in **Figure 6**. Similarly, little difference was seen between when the ELPS is activated and when it was deactivated. The difference between the highest measured quantitative T2 values against the reference values was slightly higher when the ELPS is activated, however the difference is minimal.

**Figure 6.**
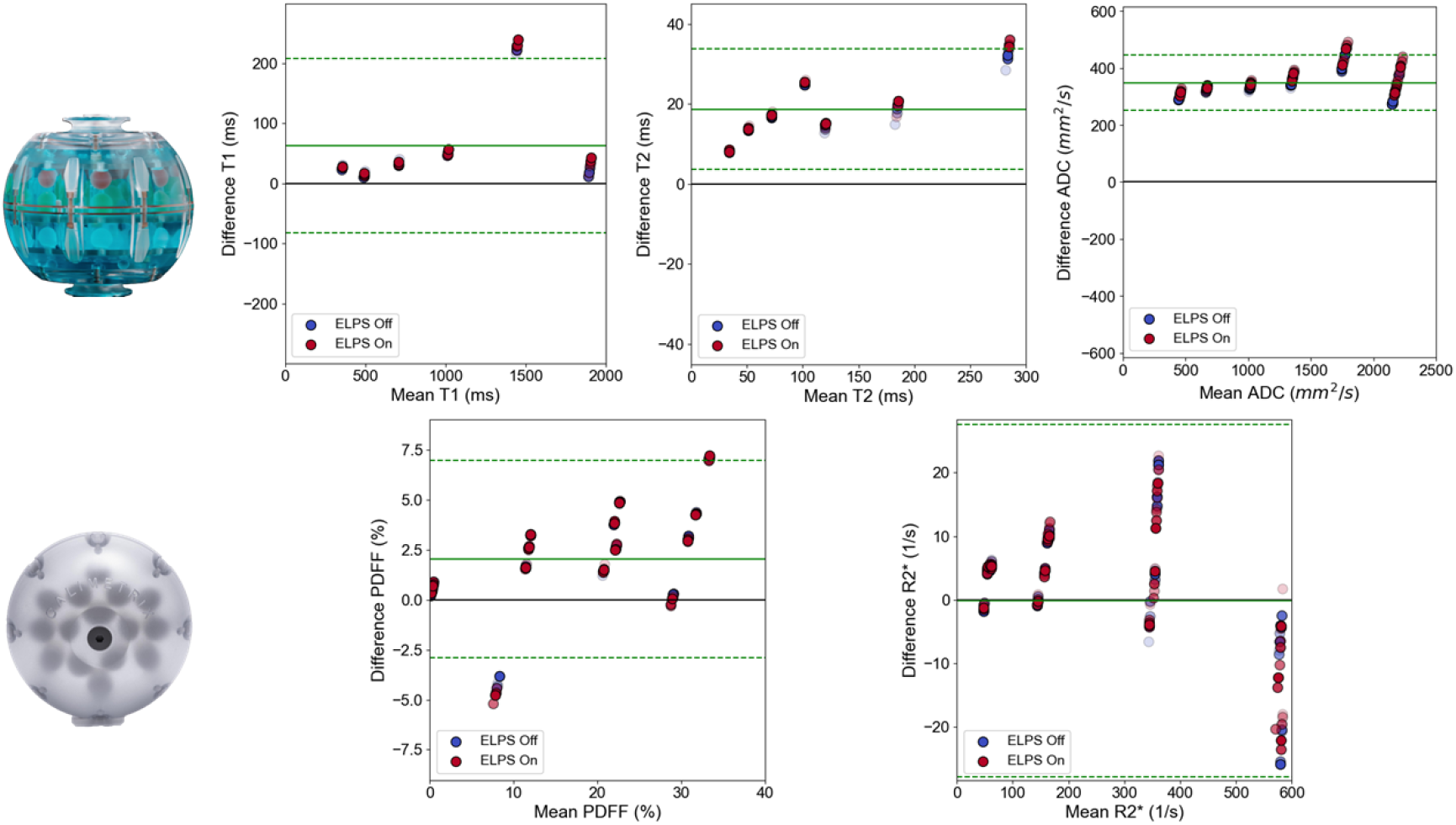
Bland-Altman plots of the quantitative T1 (top left), T2 (top middle), ADC (top right), PDFF (bottom left), and R2* (bottom right) with corresponding mean (solid green) and 95% lines of agreement (dashed green) for when the ELPS is both deactivated (blue) and activated (red).

The largest difference between when the ELPS was activated compared to when it was deactivated is shown in **Figure 7** which plots the standard deviation of the values inside each vial against their mean values. The largest difference was seen in the ADC which consistently demonstrates a higher standard deviation of the values inside the vial when the ELPS was activated.

**Figure 7.**
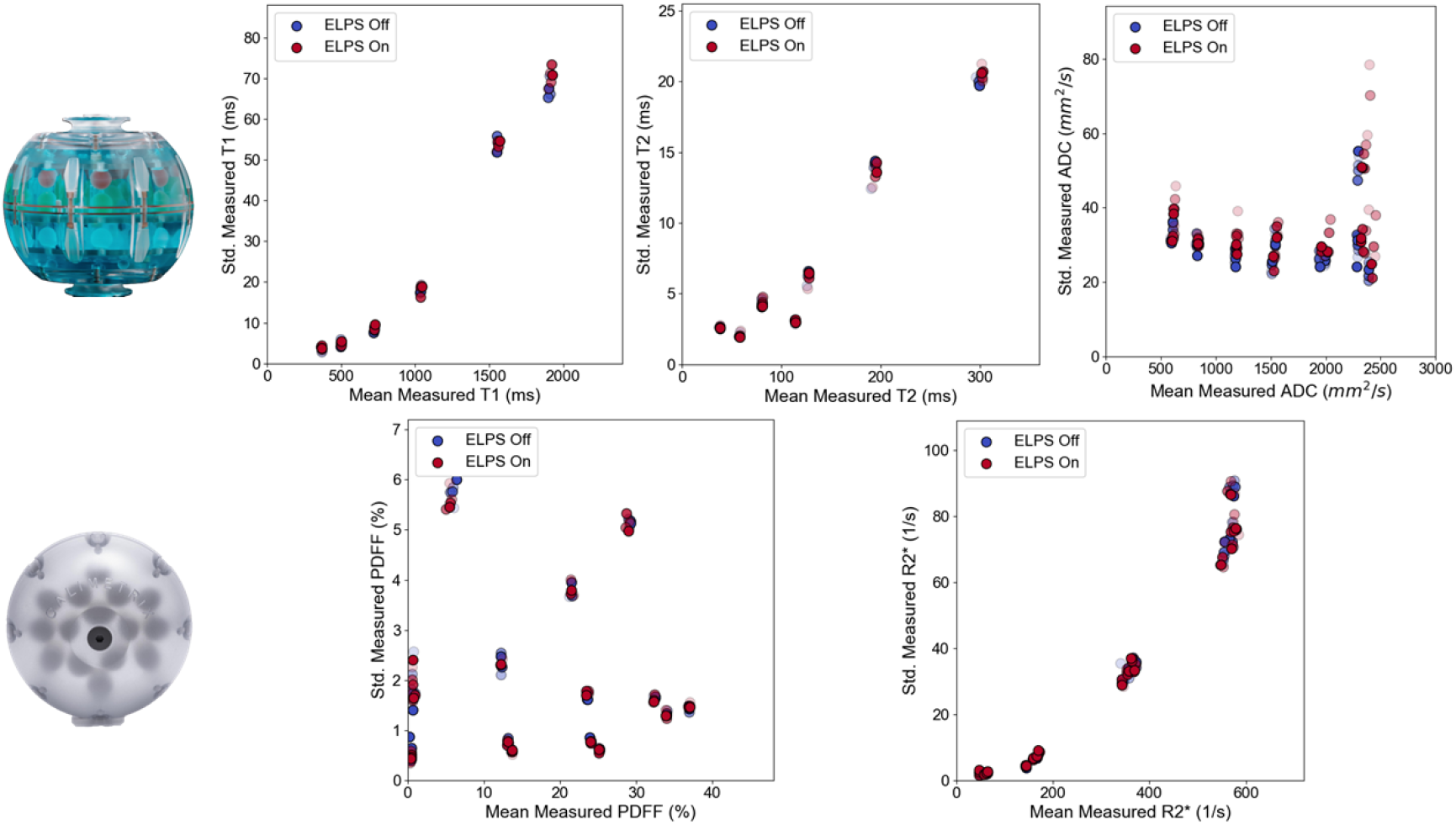
Plots of the standard deviation against the mean of the quantitative T1 (top left), T2 (top middle), ADC (top right), PDFF (bottom left), and R2* (bottom right) for when the ELPS is both deactivated (blue) and activated (red).

To visualize **Figure 7** more explicitly, **Figure 8** demonstrates how the CoV changes across the quantitative values when the ELPS is activated compared to when it was previously deactivated during the repetitions. A notable increase was seen in the ADC values when the ELPS was activated compared to when it was deactivated, especially at the higher ADC values.

**Figure 8.**
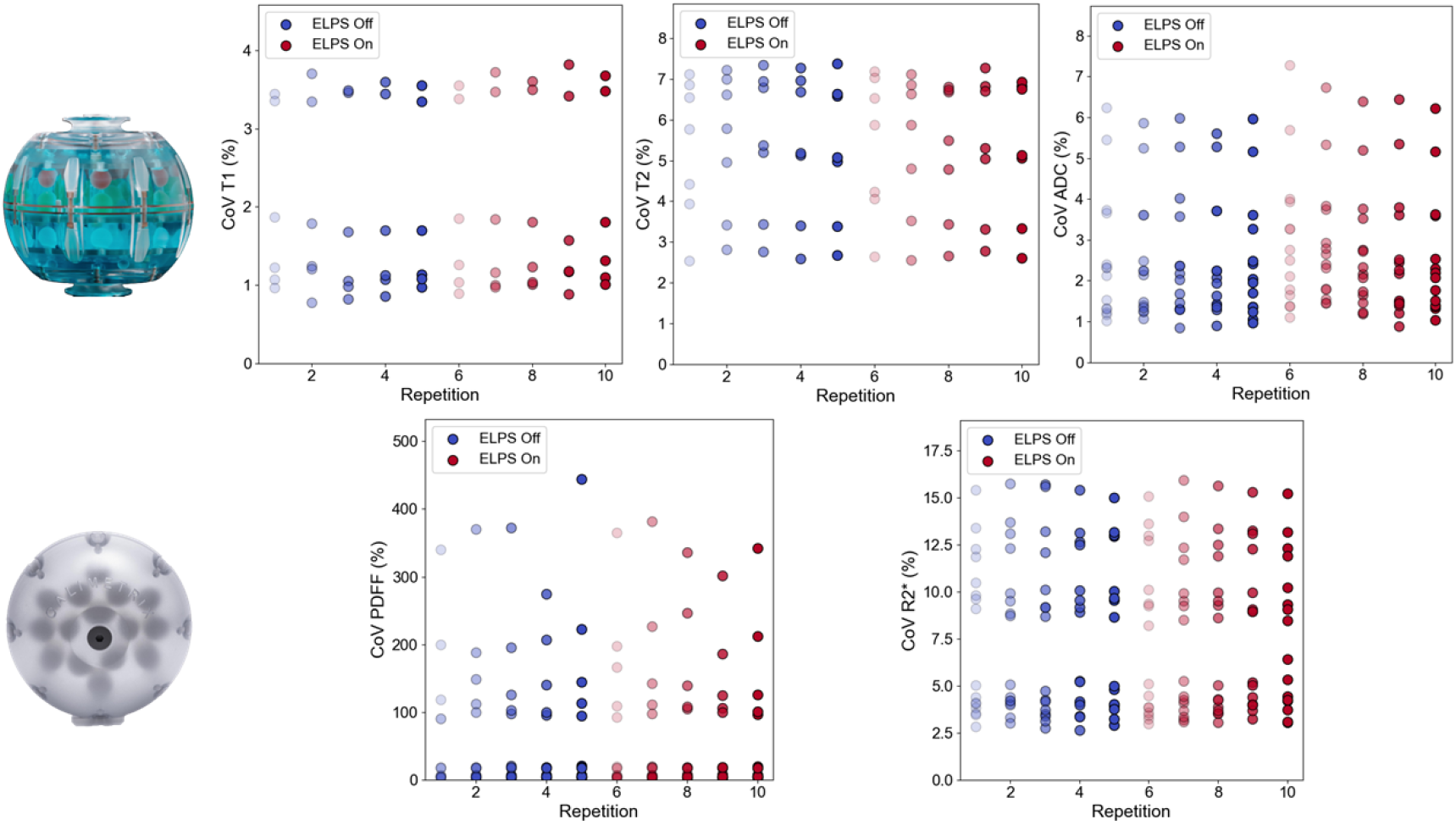
Plots of the CoV against the acquisition repetition for the quantitative T1 (top left), T2 (top middle), ADC (top right), PDFF (bottom left), and R2* (bottom right) for when the ELPS is both deactivated (blue) and activated (red).

To investigate the increased standard deviation inside the phantom vials in the BLADE DWI sequence, the SNR difference between the individual b-value images was determined to investigate the cause of this further. As shown in **Table 5**, there were minimal SNR differences among the two b-value images from when the ELPS is deactivated and activated.

**Table 5.** The change in SNR across the individual b-value images from the BLADE DWI sequence when the ELPS is both deactivated and activated.

|  | SNR<br>(ELPS Deactivated) | SNR<br>(ELPS Activated) |
| --- | --- | --- |
| b-value = 0 s/mm<sup>2</sup> | 77 | 75 |
| b-value = 800 s/mm<sup>2</sup> | 26 | 26 |

## 4. Discussion

### 4.1. Summary

This study presented the effects of ELPS activation during MRI scanning on the resulting image quality across several coil combinations and quantification accuracy and repeatability of quantitative MRI sequences for T1, T2, ADC, PDFF, and R2* using clinically used coil combinations. This is the first known study to the author’s knowledge to present such an analysis in detail. In summary, the novel contributions of this study to the characterization of ELPS activation during MRI scanning are: 1) the description of the SNR changes across different coil combinations, 2) the real-time identification of the induced electronic noise artifact using a cine acquisition, and 3) the resulting impacts on quantitative MRI accuracy and repeatability.

From the ACR localizer analysis, the largest difference in SNR was in the integrated body coil due to the lowest SNR from the individual receive elements which gets increased with the higher channel coils such as the 18-channel UltraFlex series and the dedicated anatomical coils (i.e., shoulder). This was most likely a contributing factor as to why there was little reported difference in quantitative values between when the ELPS deactivated and activated. However, degradation in image intensity uniformity, percent signal ghosting, and low contrast object detectability was seen during ACR Large Phantom testing using the 20-channel Head/Neck coil. An additional consequence shown in this study of keeping the ELPS activated during imaging is that it impacts the robustness of automated geometric distortion algorithms due to the addition of electronic noise-induced banding artifacts, contributing to localized image noise, hindering edge detection.

The largest difference in the quantitative MRI sequences was seen in the ADC, which consistently demonstrated a higher standard deviation of the values inside the vial when the ELPS was activated. Since there was little difference in the SNR measurements among the individual b-values images between when the ELPS was deactivated and activated, the difference in standard deviation may be due to noise propagation in specific vials affected the most. However, it should be noted that the difference in the CoV was also small at around 1% from 6% when the ELPS was deactivated to 7% when it was activated. Possible explanations for the high level of agreement with both the MDME quantitative T1 and T2 maps are their acquisition parameter of a GRAPPA factor of 3 which has been shown to reduce noise in some areas while increasing noise in others^26,27^. This effect could effectively nullify, or smear out, any association with electronic-induced noise from the ELPS. However, the quantitative performance of the q-Dixon sequence for PDFF and R2* was not affected even though it did not use any image acceleration techniques.

### 4.2. Limitations

The Calimetrix phantom used in this study has a maximum PDFF value of around 40% which limits its evaluation of fatty structures which are of interest in several anatomical sites such as in the head and neck where parotid salivary glands may exceed this value^28^. However, it should be noted that quantitative Dixon reconstruction algorithms should perform similarly at PDFF values symmetric around 50% due to the same fraction^29^ (i.e., 20% vs. 80%). In this study, we found a systematic positive ADC bias of ∼320 mm^2^/s which we suscept is due to two possible causes: (1) our use of b-values of 0 s/mm^2^ which includes perfusion components^30^ and (2) our use of BLADE DWI compared to a conventional echo planer imaging (EPI) technique which can lead to additional bias^31^. We determined that this bias was not due to incorrect temperature readings since the same value was used for the quantitative T1 and T2 values which did not experience a similar bias. Additionally, our SNR calculation for the BLADE DWI individual b-value images was done by using the CaliberMRI MiniHybrid phantom which does not contain a true homogeneous slice like the homogeneous phantom used in the study for the other acquisitions. Some of our SNR calculations were performed using the ACR localizer images in some cases instead of the standard T1- or T2-weighted sequence, the reported SNR values may not accurately reflect the true changes in magnitude. However, we believe that the final trends should be the same due to the extra noise induced by ELPS activation.

### 4.3. Future Work

From a technical perspective, future studies should be done to characterize the ELPS-induced artifact across different MRI system static magnetic field strengths and vendors. Our institution has a 1.5T Siemens MR-simulation scanner, however we did not notice any noticeable level of ELPS-induced artifact on that system since the ELPS system was nearly twice as far away from the MRI due to its increased room size. If true, this may be one of many considerations when site planning a new MRI system. Additionally, more robust image SNR analysis should be conducted to more accurately quantify the resulting SNR changes. It would be important to also evaluate the difference in a human subject with clinically used coil combinations, though from our results in phantoms, it should be unlikely to see a very large difference.

From an administrative and clinical workflow standpoint, the work shown here is motivational to persuade clinical staff with frequent opportunities to scan on an MRI-simulation scanner (i.e., MRI technologists, physicists, etc.) to inform and remind their colleagues of the importance of deactivating the ELPS during scanning. Additionally, clinical supervisors should consider effective ways to properly educate clinical staff of the effects of the ELPS on the resulting MRI images, with their small but, importantly, preventable addition to the inherent noise in any image. It may be of interest to determine automatic ways to deactivate the ELPS during scanning such as how autoinjectors work during dynamic contrast-enhanced (DCE) MRI examinations. Although the ELPS used in this study is MR Conditional dependent on being deactivated during MRI examination, there was a communication gap in our clinical workflow which prevented its deactivation during quality assurance testing. This recommendation from the vendor should therefore be properly carried forward to staff using its products.

## 5. Conclusion

This study demonstrated the effects of ELPS-induced electronic noise interference on MRI image quality and quantification for T1, T2, ADC, PDFF, and R2* values. Although the integrated body coil demonstrated the most noticeable reduction in image quality, even affecting fiducial detection in a geometric distortion phantom, efforts should be made from the clinical team to ensure its deactivation during imaging to eliminate preventable reduction in SNR. Finally, although ELPS activation did not severely impact mean quantitative MRI sequence results, its CoV may be slightly affected, further reinforcing the need for the clinical team to address this issue. It should be reiterated that the entire ELPS must be deactivated instead of simply turning the visual laser to eliminate the production of the RF interference leading to the imaging artifacts.

## Data Availability

All relevant image files and contours in NIfTI format are to be made publicly available after manuscript acceptance at the following DOI: 10.6084/m9.figshare.31558963. The accompanying code for image visualization and statistical analysis will be made publicly available at the following URL: https://github.com/Lucas-Mc/MRI-ELPS.

https://doi.org/10.6084/m9.figshare.31558963

https://github.com/Lucas-Mc/MRI-ELPS

## Acknowledgements

None.

## References

1. Glide-Hurst CK, Wen N, Hearshen D, et al. Initial clinical experience with a radiation oncology dedicated open 1.0T MR-simulation. J Appl Clin Med Phys. 2015;16(2):218–240. doi:10.1120/jacmp.v16i2.5201

2. Kwok WE. Radiofrequency interference in magnetic resonance imaging: Identification and rectification. J Clin Imaging Sci. 2024;14:33. doi:10.25259/JCIS_74_2024

3. Kwok WE. Radiofrequency interference in magnetic resonance imaging: Identification and rectification. J Clin Imaging Sci. 2024;14. doi:10.25259/JCIS_74_2024

4. Graves MJ, Mitchell DG. Body MRI artifacts in clinical practice: A physicist’s and radiologist’s perspective. Journal of Magnetic Resonance Imaging. 2013;38(2):269–287. doi:10.1002/jmri.24288

5. McCullum L, Mulder SL, West NA, et al. Technical development and In Silico implementation of SyntheticMR in head and neck adaptive radiation therapy: A prospective R-IDEAL stage 0/1 technology development report. Journal of Applied Clinical Medical Physics. 2025;26(7):e70134. doi:10.1002/acm2.70134

6. McDonald BA, Salzillo T, Mulder S, et al. Prospective evaluation of in vivo and phantom repeatability and reproducibility of diffusion-weighted MRI sequences on 1.5 T MRI-linear accelerator (MR-Linac) and MR simulator devices for head and neck cancers. Radiotherapy and Oncology. 2023;185. doi:10.1016/j.radonc.2023.109717

7. Costelloe CM, Madewell JE, Kundra V, Harrell RK, Bassett RL, Ma J. Conspicuity of bone metastases on fast Dixon-based multisequence whole-body MRI: Clinical utility per sequence. Magnetic Resonance Imaging. 2013;31(5):669–675. doi:10.1016/j.mri.2012.10.017

8. Clarke CN, Choi H, Hou P, et al. Using MRI to non-invasively and accurately quantify preoperative hepatic steatosis. HPB. 2017;19(8):706–712. doi:10.1016/j.hpb.2017.04.009

9. Zhao R, Hamilton G, Brittain JH, Reeder SB, Hernando D. Design and evaluation of quantitative MRI phantoms to mimic the simultaneous presence of fat, iron, and fibrosis in the liver. Magnetic Resonance in Med. 2021;85(2):734–747. doi:10.1002/mrm.28452

10. Carr HY. Steady-State Free Precession in Nuclear Magnetic Resonance. Phys Rev. 1958;112(5):1693–1701. doi:10.1103/PhysRev.112.1693

11. Bieri O, Scheffler K. Fundamentals of balanced steady state free precession MRI: Fundamentals of Balanced SSFP MRI. J Magn Reson Imaging. 2013;38(1):2–11. doi:10.1002/jmri.24163

12. Griswold MA, Jakob PM, Heidemann RM, et al. Generalized autocalibrating partially parallel acquisitions (GRAPPA). Magnetic Resonance in Med. 2002;47(6):1202–1210. doi:10.1002/mrm.10171

13. Frahm J, Haase A, Matthaei D. Rapid Three-Dimensional MR Imaging Using the FLASH Technique. Journal of Computer Assisted Tomography. 1986;10(2):363.

14. Warntjes JBM, Leinhard OD, West J, Lundberg P. Rapid magnetic resonance quantification on the brain: Optimization for clinical usage. Magnetic Resonance in Med. 2008;60(2):320–329. doi:10.1002/mrm.21635

15. Pipe JG, Farthing VG, Forbes KP. Multishot diffusion-weighted FSE using PROPELLER MRI. Magnetic Resonance in Medicine. 2002;47(1):42–52. doi:10.1002/mrm.10014

16. Lang TA, Altman DG. Statistical Analyses and Methods in the Published Literature: The SAMPL Guidelines*. In: Moher D, Altman DG, Schulz KF, Simera I, Wager E, eds. Guidelines for Reporting Health Research: A User’s Manual. 1st ed. Wiley; 2014:264–274. doi:10.1002/9781118715598.ch25

17. Kaufman L, Kramer DM, Crooks LE, Ortendahl DA. Measuring signal-to-noise ratios in MR imaging. Radiology. 1989;173(1):265–267. doi:10.1148/radiology.173.1.2781018

18. Schneider CA, Rasband WS, Eliceiri KW. NIH Image to ImageJ: 25 years of image analysis. Nat Methods. 2012;9(7):671–675. doi:10.1038/nmeth.2089

19. Fedorov A, Beichel R, Kalpathy-Cramer J, et al. 3D Slicer as an Image Computing Platform for the Quantitative Imaging Network. Magn Reson Imaging. 2012;30(9):1323–1341. doi:10.1016/j.mri.2012.05.001

20. Cox RW, Ashburner J, Breman H, et al. A (Sort of) New Image Data Format Standard: NiFTI-1. Vol 22.; 2004.

21. Hough PVC. Man - Machine Collaboration in the Analysis of Bubble Chamber Photography for High - Energy Physics. Opt Eng. 1964;2(3). doi:10.1117/12.7971269

22. Duda RO, Hart PE. Use of the Hough transformation to detect lines and curves in pictures. Commun ACM. 1972;15(1):11–15. doi:10.1145/361237.361242

23. Hart P. How the Hough transform was invented [DSP History. IEEE Signal Process Mag. 2009;26(6):18–22. doi:10.1109/MSP.2009.934181

24. Lin LIK. A Concordance Correlation Coefficient to Evaluate Reproducibility. Biometrics. 1989;45(1):255. doi:10.2307/2532051

25. Altman DG, Bland JM. Measurement in Medicine: The Analysis of Method Comparison Studies. The Statistician. 1983;32(3):307. doi:10.2307/2987937

26. Jaspan ON, Fleysher R, Lipton ML. Compressed sensing MRI: a review of the clinical literature. Br J Radiol. 2015;88(1056):20150487. doi:10.1259/bjr.20150487

27. Robson PM, Grant AK, Madhuranthakam AJ, Lattanzi R, Sodickson DK, McKenzie CA. Comprehensive Quantification of Signal-to-Noise Ratio and g-Factor for Image-Based and k-Space-Based Parallel Imaging Reconstructions. Magn Reson Med. 2008;60(4):895–907. doi:10.1002/mrm.21728

28. Chikui T, Yamashita Y, Kise Y, Saito T, Okamura K, Yoshiura K. Estimation of proton density fat fraction of the salivary gland. Br J Radiol. 2018;91(1085):20170671. doi:10.1259/bjr.20170671

29. Daudé P, Roussel T, Troalen T, et al. Comparative review of algorithms and methods for chemical-shift-encoded quantitative fat-water imaging. Magnetic Resonance in Medicine. 2024;91(2):741–759. doi:10.1002/mrm.29860

30. Le Bihan D. What can we see with IVIM MRI? NeuroImage. 2019;187:56–67. doi:10.1016/j.neuroimage.2017.12.062

31. McDonald BA, Salzillo T, Mulder S, et al. Prospective evaluation of in vivo and phantom repeatability and reproducibility of diffusion-weighted MRI sequences on 1.5 T MRI-linear accelerator (MR-Linac) and MR simulator devices for head and neck cancers. Radiotherapy and Oncology. 2023;185. doi:10.1016/j.radonc.2023.109717

